# Antibody prevalence for SARS-CoV-2 following the peak of the pandemic in England: REACT2 study in 100,000 adults

**DOI:** 10.1101/2020.08.12.20173690

**Authors:** Helen Ward, Christina Atchison, Matthew Whitaker, Kylie EC Ainslie, Joshua Elliott, Lucy Okell, Rozlyn Redd, Deborah Ashby, Christl A Donnelly, Wendy Barclay, Ara Darzi, Graham Cooke, Steven Riley, Paul Elliott

**Affiliations:** School of Public Health, Imperial College London; Imperial College Healthcare NHS Trust; National Institute for Health Research Imperial Biomedical Research Centre; School of Public Health, Imperial College London; Imperial College Healthcare NHS Trust; School of Public Health, Imperial College London; School of Public Health, Imperial College London; MRC Centre for Global Infectious Disease Analysis Imperial College London; School of Public Health, Imperial College London; MRC Centre for Global Infectious Disease Analysis, Imperial College London; Department of Statistics, University of Oxford; Department of Infectious Disease, Imperial College London; Imperial College Healthcare NHS Trust; Institute of Global Health Innovation, Imperial College London; Imperial College Healthcare NHS Trust; Department of Infectious Disease, Imperial College London; Imperial College Healthcare NHS Trust; National Institute for Health Research Imperial Biomedical Research Centre; School of Public Health, Imperial College London; MRC Centre for Global Infectious Disease Analysis, Imperial College London; School of Public Health, Imperial College London; Imperial College Healthcare NHS Trust; National Institute for Health Research Imperial Biomedical Research Centre; MRC Centre for Environment and Health, Imperial College London

**Author notes:** **CORRESPONDING AUTHOR** Professor Paul Elliott, Address: School of Public Health, Imperial College London, London W2 1PG, UK.

## Abstract

**Background:** England, UK has experienced a large outbreak of SARS-CoV-2 infection. As in USA and elsewhere, disadvantaged communities have been disproportionately affected.

**Methods:** National REal-time Assessment of Community Transmission-2 (REACT-2) prevalence study using a self-administered lateral flow immunoassay (LFIA) test for IgG among a random population sample of 100,000 adults over 18 years in England, 20 June to 13 July 2020.

**Results:** Data were available for 109,076 participants, yielding 5,544 IgG positive results; adjusted (for test performance) and re-weighted (for sampling) prevalence was 6.0% (95% CI: 5.8, 6.1). Highest prevalence was in London (13.0% [12.3, 13.6]), among people of Black or Asian (mainly South Asian) ethnicity (17.3% [15.8, 19.1] and 11.9% [11.0, 12.8] respectively) and those aged 18-24 years (7.9% [7.3, 8.5]). Adjusted odds ratio for care home workers with client-facing roles was 3.1 (2.5, 3.8) compared with non-essential workers. One third (32.2%, [31.0-33.4]) of antibody positive individuals reported no symptoms. Among symptomatic cases, most (78.8%) reported symptoms during the peak of the epidemic in England in March (31.3%) and April (47.5%) 2020. We estimate that 3.36 million (3.21, 3.51) people have been infected with SARS-CoV-2 in England to end June 2020, with an overall infection fatality ratio (IFR) of 0.90% (0.86, 0.94); age-specific IFR was similar among people of different ethnicities.

**Conclusion:** The SARS-CoV-2 pandemic in England disproportionately affected ethnic minority groups and health and care home workers. The higher risk of infection in minority ethnic groups may explain their increased risk of hospitalisation and mortality from COVID-19.

## Introduction

England has experienced a large outbreak of SARS-CoV-2 infection leading to the highest excess mortality in Europe by June 2020.(1) The first COVID-19 death occurred on 28 February, with in-hospital deaths peaking at 800 per day within 6 weeks.(2) Hospital admission and mortality data show an asymmetrical burden of COVID-19 in England, with high rates in older people including those living in long-term care, and in people of minority ethnic groups, particularly Black and Asian (mainly South Asian) individuals.(3-6) It is unclear how much of this excess is due to differences in exposure to the virus, e.g. related to workplace exposures and structural inequality, and how much is due to differences in outcome, including access to health care.(7, 8)

Antibody data provide a long-lasting measure of SARS-CoV-2 infection, enabling analyses of the recent epidemic. Most infected people mount an IgG antibody response detectable after 14-21 days although levels may start to wane after ~90 days.(9,10) Uncertain validity of the available antibody tests, inconsistencies in sampling methods, small numbers and use of selected groups have made many studies difficult to interpret.(11) Different criteria may apply to community-based compared with individual studies where population-wide results are required.(10-15) Self-administered lateral flow immunoassay (LFIA) tests done at home offer a method for obtaining community-wide prevalence estimates rapidly and at scale, at reasonable cost. While there have been questions about their use for individual care,(16-18) reliable population prevalence estimates can be obtained by adjusting for known test performance.(19)

We aimed here to i) estimate the cumulative community prevalence of IgG antibodies for SARS-CoV-2 from a large representative sample in England up to early July 2020, ii) identify those at most risk of infection, and iii) estimate the total number of infected individuals in England as well as the infection fatality ratio (IFR).

## Methods

The REal-time Assessment of Community Transmission-2 (REACT-2) programme is evaluating community prevalence of SARS-CoV-2 infection in England.(20) We obtained a random population sample of adults in England, using the National Health Service (NHS) patient list, which includes name, address, age and sex of everyone registered with a general practitioner (almost the entire population). Personalized invitations were sent to 315,000 individuals aged 18 years and above to achieve similar numbers in each of 315 lower-tier local authority areas. Participants registered via an online portal or by telephone with registration closed after ~120,000 people had signed up.

Those registered were sent a test kit, including a self-administered point-of-care LFIA test and instructions by post, with link to an on-line video. The LFIA (Fortress Diagnostics, Northern Ireland) was selected following evaluation of performance characteristics (sensitivity and specificity) against pre-defined criteria for detection of IgG,(21) and extensive public involvement and user testing.(22) Compared to results from at least one of two in house ELISAs, sensitivity and specificity of finger-prick blood (self-read) were 84.4% (70.5, 93.5) in RT-PCR confirmed cases and 98.6% (97.1, 99.4) in 500 pre-pandemic sera.(21) Participants completed a short registration questionnaire (online/telephone) and a further survey upon completion of their self-test. This included information on demographics, household composition, recent symptoms and an uploaded photograph of the result. A validation study of the photographs showed substantial concordance between participant- and clinician-interpreted results in over 500 tests (kappa: 0.89, 95% Cl: 0.88-0.92).(22)

Prevalence was calculated as the proportion of individuals with a positive IgG result, adjusted for test performance using:

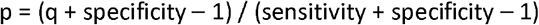

where p = adjusted proportion positive, q = observed proportion positive.(19) Prevalence estimates at national level were weighted for age, sex, region, ethnicity and deprivation to account for the geographic sample design and for variation in response rates, so as to be representative of the population (18+ years) of England. Logistic regression models were adjusted for age, sex and region, and additionally for ethnicity, deprivation, household size and occupation. We used complete case analysis without imputation.

We estimated total number of SARS-CoV-2 infections since start of the epidemic until July 2020 by multiplying the antibody prevalence, adjusted for test characteristics and re-weighted for sampling, by mid-year population size at ages 18+ years in England.(23) To correct for survival bias we added to the seropositive population the deaths that mentioned COVID-19 on the death certificate during this period. We then estimated the IFR, dividing the total number of COVID-19 deaths excluding care home residents.(14) We obtained an overall IFR estimate and estimates stratified by age and gender.(24) Confidence bounds were obtained by using the Delta method. As a sensitivity analysis we calculated IFR and total infections including care home residents and with all-cause excess deaths and stratified by ethnicity, age and sex.

We obtained research ethics approval from the South Central-Berkshire B Research Ethics Committee (IRAS ID: 283787), and MHRA approval for use of the LFIA for research purposes only.

Data were analysed using the statistical package R version 4.0.0.(25)

## Results

Of the 121,976 people who were sent LFIA test kits, 109,076 (89.4%) completed the questionnaire of whom 105,651 also completed the test; 5,743 (5.4%) reported an invalid or unreadable result leaving 99,908 (94.6%) individuals, 5,544 IgG positive and 94,364 IgG negative, giving a crude prevalence of 5.6% (95% CI 5.4-5.7). After adjusting for the performance characteristics of the test and re-weighting, overall prevalence for England was 6.0% (95% Cl: 5.8-6.1) during the period 20 June to 13 July 2020. This equates to 3.36 (3.22, 3.51) million adults in England who had been infected with SARS-CoV-2 in England to end June 2020.

Prevalence was highest for ages 18-24 years and in London (Table 1). Highest prevalence by ethnic group was found in people of Black (includes Black Caribbean, African and Black British) (17.3%, 95% CI 15.8,19.1) and Asian (mainly South Asian) ethnicities (11.9%, 95% CI 11.0,12.8), compared to 5.0% (4.8, 5.2) in people of white ethnicity (Table 1). The association of prevalence with non-white ethnicities was partially but not fully explained by the covariates. For example, in the unadjusted model, compared to white ethnicity, Black ethnicity was associated with a three-fold increase in odds of being antibody positive (OR 3.2, 95%CI 2.7, 3.9) which reduced to OR 2.0 (1.6, 2.4) after adjustment for covariates (Figure 1, Supplementary Appendix Table S1). Essential workers, particularly those with public-facing roles, also had increased prevalence. Prevalence among those working in residential care facilities (care homes) with client-facing roles was 16.5% (95% CI 13.7, 19.8) and it was 11.7% (95% CI 10.5-13.1) among health care workers with patient contact, with 3-fold (3.09; 2.51,3.80) and 2-fold (2.09; 1.86,2.35) odds of infection respectively compared with non-essential workers (Table 2, Figure 1). Those in the more deprived areas or living in larger households had higher prevalence than those in more affluent areas or who lived alone, although the increased odds were partially attenuated in the adjusted models; people who currently smoked had a lower prevalence (3.2%, 2.8, 3.7) than those who did not (5.2%, 5.0, 5.4), OR 0.64 [0.58,0.71) (Table 2, Figure 1, Supplementary Appendix Table S1).

**Table 1:** Prevalence of SARS-CoV-2 IgG antibodies: crude, adjusted for test performance, and weighted to the population (18+ years) of England, by sociodemographic characteristics.

|  | *Total*<br>*antibody*<br>*positive* | *Total tests*<br>*(with valid*<br>*results)* | *Crude prevalence*<br>*%*<br>*[95% confidence*<br>*intervals]* | *Prevalence %*<br>*adjusted for test*<br>*[95% confidence*<br>*intervals]* | *Weighted<sup>1</sup>*<br>*prevalence %*<br>*[95% confidence*<br>*intervals]* |
| --- | --- | --- | --- | --- | --- |
| *England* | 5544 | 99908 | 5.6 [5.4-5.7] | 5.0 [4.8-5.2] | 6.0 [5.8-6.1] |
| **Sex** |  |  |  |  |  |
| *Male* | 2405 | 43825 | 5.5 [5.3-5.7] | 4.9 [4.7-5.2] | 6.2 [5.9-6.4] |
| *Female* | 3139 | 56083 | 5.6 [5.4-5.8] | 5.1 [4.8-5.3] | 5.8 [5.5-6.0] |
| **Age** |  |  |  |  |  |
| *18-24* | 463 | 6499 | 7.1 [6.5-7.8] | 6.9 [6.2-7.7] | 7.9 [7.3-8.5] |
| *25-34* | 930 | 13366 | 7.0 [6.5-7.4] | 6.7 [6.2-7.2] | 7.8 [7.4-8.3] |
| *35-44* | 964 | 17052 | 5.7 [5.3-6.0] | 5.1 [4.7-5.6] | 6.1 [5.7-6.6] |
| *45-54* | 1255 | 20634 | 6.1 [5.8-6.4] | 5.6 [5.3-6.0] | 6.4 [6.0-6.9] |
| *55-64* | 1131 | 20404 | 5.5 [5.2-5.9] | 5.0 [4.6-5.4] | 5.9 [5.5-6.4] |
| *65-74* | 568 | 15543 | 3.7 [3.4-4.0] | 2.7 [2.4-3.1] | 3.2 [2.8-3.6] |
| *75+* | 233 | 6410 | 3.6 [3.2-4.1] | 2.7 [2.2-3.3] | 3.3 [2.9-3.8] |
| **Ethnicity** |  |  |  |  |  |
| *White* | 4827 | 92737 | 5.2 [5.1-5.3] | 4.6 [4.4-4.8] | 5.0 [4.8-5.2] |
| *Mixed* | 106 | 1347 | 7.9 [6.5-9.4] | 7.8 [6.2-9.7] | 8.9 [7.1-11.1] |
| *Asian<sup>2</sup>* | 369 | 3658 | 10.1 [9.2-11.1] | 10.5 [9.3-11.7] | 11.9 [11.0-12.8] |
| *Black<sup>3</sup>* | 135 | 900 | 15.0 [12.8-17.5] | 16.4 [13.8-19.4] | 17.3 [15.8-19.0] |
| *Other* | 79 | 762 | 10.4 [8.4-12.7] | 10.8 [8.4-13.7] | 12.3 [10.2-14.7] |
| **Deprivation**<br>**Quintile<sup>4</sup>** |  |  |  |  |  |
| *1 most deprived* | 682 | 10082 | 6.8 [6.3-7.3] | 6.5 [5.9-7.1] | 7.3 [6.8-7.7] |
| *2* | 947 | 16015 | 5.9 [5.6-6.3] | 5.4 [5.0-5.9] | 6.4 [6.0-6.8] |
| *3* | 1196 | 21474 | 5.6 [5.3-5.9] | 5.0 [4.7-5.4] | 5.9 [5.5-6.3] |
| *4* | 1287 | 24840 | 5.2 [4.9-5.5] | 4.6 [4.2-4.9] | 5.2 [4.8-5.6] |
| *5 least deprived* | 1432 | 27497 | 5.2 [5.0-5.5] | 4.6 [4.3-4.9] | 5.0 [4.6-5.4] |
| **Household size** |  |  |  |  |  |
| *1* | 720 | 15052 | 4.8 [4.5-5.1] | 4.1 [3.7-4.5] | 4.7 [4.3-5.1] |
| *2* | 1784 | 36413 | 4.9 [4.7-5.1] | 4.2 [4.0-4.5] | 5.0 [4.7-5.3] |
| *3* | 1158 | 19734 | 5.9 [5.5-6.2] | 5.4 [5.0-5.8] | 6.5 [6.0-6.9] |
| *4* | 1204 | 19611 | 6.1 [5.8-6.5] | 5.7 [5.3-6.1] | 6.4 [6.0-6.8] |
| *5* | 447 | 6403 | 7.0 [6.4-7.6] | 6.7 [6.0-7.5] | 7.7 [7.0-8.5] |
| *6* | 152 | 1848 | 8.2 [7.1-9.6] | 8.2 [6.8-9.8] | 12.3 [10.8-14.0] |
| *7+* | 79 | 827 | 9.6 [7.7-11.7] | 9.8 [7.6-12.5] | 13.0 [11.0-15.3] |
| **Region** |  |  |  |  |  |
| *North East* | 196 | 3574 | 5.5 [4.8-6.3] | 4.9 [4.1-5.9] | 5.0 [4.3-5.9] |
| *North West* | 714 | 11996 | 6.0 [5.5-6.4] | 5.5 [5.0-6.0] | 6.6 [6.1-7.2] |
| *Yorkshire* | 284 | 6519 | 4.4 [3.9-4.9] | 3.6 [3.0-4.2] | 3.9 [3.5-4.5] |
| *East Midlands* | 601 | 12684 | 4.7 [4.4-5.1] | 4.0 [3.6-4.5] | 4.2 [3.7-4.8] |
| *West Midlands* | 547 | 9620 | 5.7 [5.2-6.2] | 5.2 [4.6-5.7] | 5.8 [5.3-6.4] |
| *East of England* | 805 | 14433 | 5.6 [5.2-6.0] | 5.0 [4.6-5.5] | 5.1 [4.6-5.6] |
| *London* | 1045 | 9547 | 10.9 [10.3-11.6] | 11.5 [10.8-12.3] | 13.0 [12.3-13.6] |
| *South East* | 995 | 21979 | 4.5 [4.3-4.8] | 3.8 [3.4-4.1] | 3.9 [3.5-4.3] |
| *South West* | 357 | 9556 | 3.7 [3.4-4.1] | 2.8 [2.4-3.3] | 2.8 [2.4-3.3] |

<sup>1</sup>All estimates of prevalence adjusted for imperfect test sensitivity and specificity (see text for details). Responses have been re-weighted to account for sample design and for variation in response rate (age, gender, ethnicity and deprivation) in final column to be representative of the England population (18+); <sup>2</sup> Asian / Asian British; <sup>3</sup> Black / African / Caribbean / Black British; <sup>4</sup> Based on Index of Multiple Deprivation (2019) at lower super output area.

**Table 2:** Prevalence of SARS-CoV-2 IgG antibodies: crude and adjusted for test performance, for individual and clinical characteristics.

|  | *Total antibody*<br>*positive* | *Total tests*<br>*(with valid*<br>*results)* | *Crude prevalence %*<br>*(95% CI)* | *Prevalence %*<br>*adjusted for test* |
| --- | --- | --- | --- | --- |
| **Employment** |  |  |  |  |
| *Health care (patient-facing)* | 379 | 3402 | 11.1 [10.1-12.2] | 11.7 [10.5-13.1] |
| *Health care (other)* | 73 | 1151 | 6.3 [5.1-7.9] | 6.0 [4.4-7.8] |
| *Care home (client-facing)* | 115 | 761 | 15.1 [12.7-17.8] | 16.5 [13.7-19.8] |
| *Care home (other)* | 12 | 146 | 8.2 [4.8-13.8] | 8.2 [4.1-15.0] |
| *Other essential worker<sup>1</sup>* | 1209 | 19927 | 6.1 [5.7-6.4] | 5.6 [5.2-6.0] |
| *Other worker* | 2189 | 37855 | 5.8 [5.6-6.0] | 5.3 [5.0-5.6] |
| *Not in employment* | 1516 | 35737 | 4.2 [4.0-4.5] | 3.4 [3.2-3.7] |
| **History of COVID-19** |  |  |  |  |
| *Positive PCR test* | 277 | 341 | 81.2 [76.7-85.0] | 96.2 [90.8-100.0] |
| *Suspected by doctor* | 353 | 1144 | 30.9 [28.2-33.6] | 35.5 [32.3-38.8] |
| *Suspected by respondent* | 3118 | 17893 | 17.4 [16.9-18.0] | 19.3 [18.6-20.0] |
| *No* | 1698 | 80390 | 2.1 [2.0-2.2] | 0.9 [0.7-1.0] |
| **Symptoms** |  |  |  |  |
| *Appetite loss* | 1504 | 5895 | 25.5 [24.4-26.6] | 29.1 [27.7-30.4] |
| *Nausea/vomiting* | 516 | 2286 | 22.6 [20.9-24.3] | 25.5 [23.5-27.6] |
| *Diarrhoea* | 744 | 3048 | 24.4 [22.9-26.0] | 27.7 [25.9-29.6] |
| *Abdominal pain* | 452 | 2230 | 20.3 [18.7-22.0] | 22.7 [20.8-24.8] |
| *Runny nose* | 732 | 4327 | 16.9 [15.8-18.1] | 18.7 [17.4-20.1] |
| *Sneezing* | 550 | 3293 | 16.7 [15.5-18.0] | 18.4 [16.9-20.0] |
| *Blocked nose* | 743 | 4217 | 17.6 [16.5-18.8] | 19.5 [18.2-21.0] |
| *Sore eyes* | 610 | 2945 | 20.7 [19.3-22.2] | 23.3 [21.6-25.1] |
| *Loss of sense of smell* | 2159 | 4714 | 45.8 [44.4-47.2] | 53.5 [51.8-55.2] |
| *Loss of sense of taste* | 2212 | 5247 | 42.2 [40.8-43.5] | 49.1 [47.5-50.7] |
| *Sore throat* | 1336 | 8994 | 14.9 [14.1-15.6] | 16.2 [15.3-17.1] |
| *Hoarse voice* | 502 | 3376 | 14.9 [13.7-16.1] | 16.2 [14.8-17.7] |
| *Headache* | 2128 | 10374 | 20.5 [19.7-21.3] | 23.0 [22.1-24.0] |
| *Dizziness* | 824 | 3956 | 20.8 [19.6-22.1] | 23.4 [21.9-25.0] |
| *Shortness of breath* | 1259 | 6899 | 18.2 [17.4-19.2] | 20.3 [19.2-21.4] |
| *New persistent cough* | 1537 | 9746 | 15.8 [15.1-16.5] | 17.3 [16.5-18.2] |
| *Tight chest* | 1228 | 6931 | 17.7 [16.8-18.6] | 19.7 [18.6-20.8] |
| *Chest pain* | 590 | 3408 | 17.3 [16.1-18.6] | 19.2 [17.7-20.7] |
| *Fever* | 1906 | 9128 | 20.9 [20.1-21.7] | 23.5 [22.5-24.5] |
| *Chills* | 1233 | 5329 | 23.1 [22.0-24.3] | 26.2 [24.8-27.6] |
| *Difficulty sleeping* | 930 | 5366 | 17.3 [16.3-18.4] | 19.2 [18.0-20.4] |
| *Tiredness* | 2626 | 12246 | 21.4 [20.7-22.2] | 24.1 [23.3-25.0] |
| *Severe fatigue* | 1256 | 5089 | 24.7 [23.5-25.9] | 28.0 [26.6-29.5] |
| *Numbness/tingling* | 358 | 1446 | 24.8 [22.6-27.0] | 28.1 [25.5-30.9] |
| *Heavy arms/legs* | 1173 | 5196 | 22.6 [21.5-23.7] | 25.5 [24.2-26.9] |
| *Muscle aches* | 2236 | 10205 | 21.9 [21.1-22.7] | 24.7 [23.8-25.7] |
| *None of these symptoms* | 6 | 74 | 8.1 [3.8-16.6] | 8.1 [2.9-18.3] |
| **Symptom severity** |  |  |  |  |
| *None* | 87 | 683 | 12.7 [10.4-15.4] | 13.7 [10.9-16.9] |
| *Mild symptoms* | 1025 | 6071 | 16.9 [16.0-17.8] | 18.7 [17.5-19.8] |
| *Moderate symptoms* | 1850 | 9261 | 20.0 [19.2-20.8] | 22.4 [21.4-23.4] |
| *Severe symptoms* | 884 | 3501 | 25.2 [23.8-26.7] | 28.7 [27.0-30.5] |
| **Contact with case** |  |  |  |  |
| *Yes, with confirmed case* | 742 | 3946 | 18.8 [17.6-20.1] | 21.0 [19.5-22.5] |
| *Yes, with suspected case* | 896 | 5307 | 16.9 [15.9-17.9] | 18.7 [17.5-19.9] |
| *No* | 3906 | 90655 | 4.3 [4.2-4.4] | 3.5 [3.3-3.7] |
| **Number of pre-existing health conditions<sup>2</sup>** |  |  |  |  |
| *>1* | 1097 | 22127 | 5.0 [4.7-5.3] | 4.3 [4.0-4.6] |
| *1* | 1558 | 28308 | 5.5 [5.2-5.8] | 4.9 [4.6-5.3] |
| *0* | 2889 | 49473 | 5.8 [5.6-6.0] | 5.3 [5.1-5.6] |
| **BMI<sup>3</sup>** |  |  |  |  |
| *Underweight (<18.5)* | 65 | 1236 | 5.3 [4.1-6.6] | 4.6 [3.3-6.3] |
| *Normal (18.5-24.9)* | 1919 | 36191 | 5.3 [5.1-5.5] | 4.7 [4.4-5.0] |
| *Overweight (25-29.9)* | 1826 | 31818 | 5.7 [5.5-6.0] | 5.2 [4.9-5.5] |
| *Obese (≥30)* | 1161 | 19855 | 5.8 [5.5-6.2] | 5.4 [5.0-5.8] |
| **Smoking (current)** |  |  |  |  |
| *Yes* | 433 | 10635 | 4.1 [3.7-4.5] | 3.2 [2.8-3.7] |
| *No* | 5059 | 88290 | 5.7 [5.6-5.9] | 5.2 [5.0-5.4] |
| **Care home resident** |  |  |  |  |
| *Yes* | 6 | 131 | 4.6 [2.1-9.6] | 3.8 [0.9-9.9] |

\* All estimates of prevalence adjusted for imperfect test sensitivity and specificity. <sup>1</sup> List of essential workers from UK government https://www.gov.uk/guidance/coronavirus-covid-19-getting-tested#essential-workers. <sup>2</sup> Pre-existing health conditions included: organ transplant recipient, diabetes (type I or II), heart disease or heart problems, hypertension, overweight, stroke, kidney disease, liver disease, anemia, asthma, other lung condition, cancer, condition affecting the brain and nerves, weakened immune system/reduced ability to deal with infections, depression, anxiety, psychiatric disorder, none of these; <sup>3</sup> Body Mass Index (BMI) formula: weight (kg) / (height [m])<sup>2</sup>.

**Figure 1:**
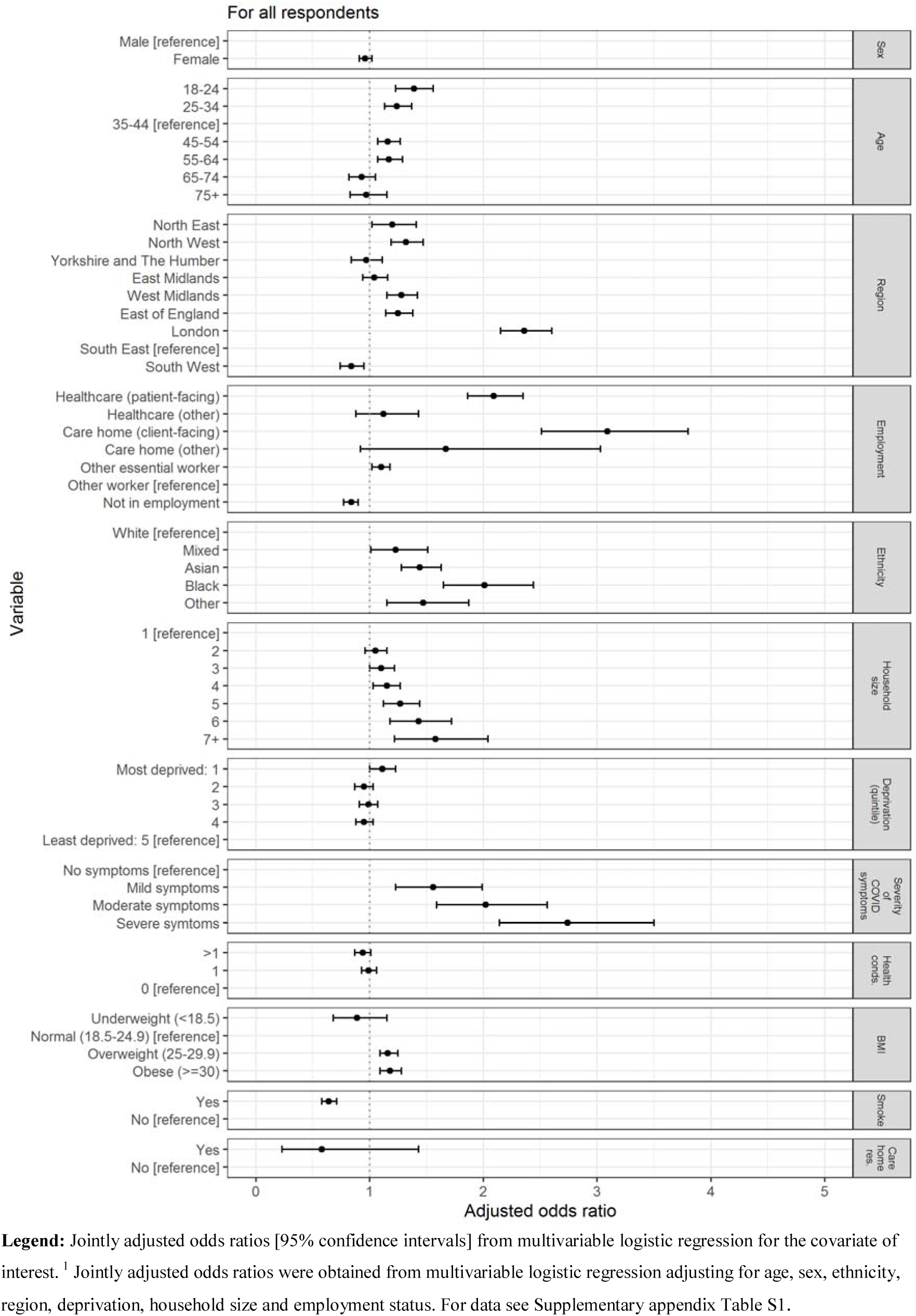
Logistic regression for SARS-CoV-2 antibodies: adjusted Odds Ratios (95% CI) for sociodemographic and clinical covariates. **Legend:** Jointly adjusted odds ratios [95% confidence intervals] from multivariable logistic regression for the covariate of interest. ^1^ Jointly adjusted odds ratios were obtained from multivariable logistic regression adjusting for age, sex, ethnicity, region, deprivation, household size and employment status. For data see Supplementary appendix Table S1.

Figure 2 shows how the epidemic evolved between January and June 2020. An epidemic curve was generated from dates of reported suspected or confirmed COVID-19 among symptomatic cases with antibodies (n=3,493) (asymptomatic individuals and symptomatic people whose date of infection was unknown are excluded). The top left plot (A) shows the epidemic curve fom the present study alongside national mortality for England by date of death - this tracks 2-3 weeks behind our epidemic curve, which peaked in the first week of April. The other panels show the proportionate distribution of cases from our data by (B) region, (C) ethnicity and (D) employment. The epidemic was widely distributed across regions; as the epidemic grew there was a shift towards a greater proportion of cases in minority ethnic groups, and in essential workers, particularly those in people-facing roles in care homes and health care.

**Figure 2.**
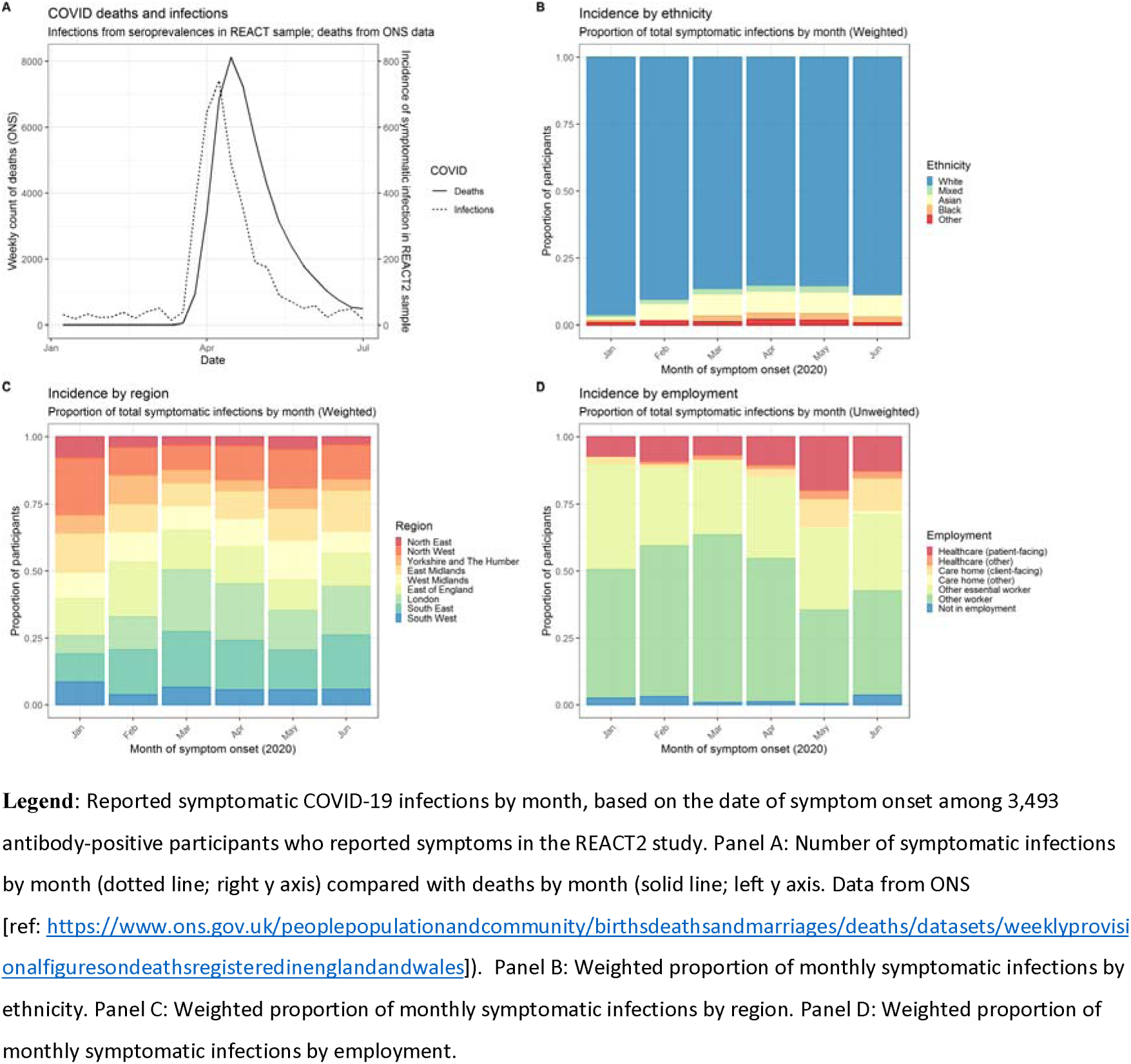
COVID-19 epidemic in England, January to June 2020, based on participants with SARS-CoV-2 antibodies by date of confirmed or suspected COVID-19 (A) compared to COVID deaths in England; and showing proportionate distribution of cases over time by (B) ethnicity (C) region (D) employment. **Legend:** Reported symptomatic COVID-19 infections by month, based on the date of symptom onset among 3,493 antibody-positive participants who reported symptoms in the REACT2 study. Panel A: Number of symptomatic infections by month (dotted line; right y axis) compared with deaths by month (solid line; left y axis. Data from ONS [ref: https://www.ons.gov.uk/peoplepopulationandcommunitv/birthsdeathsandmarriaees/deaths/datasets/weeklvprovisionalfiguresondeathsregisteredinenglandandwales]). Panel B: Weighted proportion of monthly symptomatic infections by ethnicity. Panel C: Weighted proportion of monthly symptomatic infections by region. Panel D: Weighted proportion of monthly symptomatic infections by employment.

### Clinical presentation

Of the 5,544 IgG positive people, 3,406 (61.4%; 60.1, 62.7) reported one or more typical symptoms (fever, persistent cough, loss of taste or smell), 353 (6.4%; 5.8, 7.0) reported atypical symptoms only, and 1,785 (32.2%; 31.0, 33.4) reported no symptoms. This varied by age, with people over 65 being more likely to report no symptoms (392/801, 48.9%, 45.4, 52.4) than those aged 18-34 (418/1,393, 30.0%, 27.6, 32.4) or 35-64 years (975/3,350, 29.1%, 27.6, 30.6), (p<0.001). Prevalence was higher in those with more severe symptoms, and who had contact with a confirmed or suspected case; those who were overweight or obese had higher prevalence than those with normal weight, and current smokers had a lower prevelance than non-smokers (3.2% vs. 5.2% (OR 0.64 [0.58,0.71]) (Table 2, Figure 1, Supplementary Appendix Table S1).

### Infection Fatality Ratio

The estimated community IFR (excluding care homes) was 0.90% (0.86, 0.94). It was higher in males (1.07%, 1.00,1.15) than females (0.71%, 0.67, 0.75) and increased with age from 0.52% (0.49,0.55) at ages 45-64 years to 11.64% (9.22, 14.06) at ages 75+ years (Table 3). Sensitivity analyses indicate an IFR as high as 1.58% (1.51%, 1.65%) if excess rather than COVID-specific deaths are used and care home deaths are included (Supplementary Appendix Table S2a). The estimated IFR was similar for people of Black, Asian and white ethnicities when stratified by age and sex (Supplementary Appendix Table S2b).

**Table 3:** Infection Fatality Ratio and estimated total numbers of infections.

| *Category* | *Population size, thousands* | *SARS-CoV-2 antibody prevalence % (95% CI)<sup>1</sup>* | *Confirmed COVID-19 deaths\** | *Infection fatality ratio % (95% CI)<sup>2</sup>* | *Estimated number of infections thousands (95% CI)* |
| --- | --- | --- | --- | --- | --- |
| **Total** | 56,287 | 5.96% (5.70%, 6.75%) | 30180 | 0.90% (0.86%, 0.94%) | 3,362 (3,217; 3,507) |
| **Sex** |  |  |  |  |  |
| Male | 27,828 | 6.17% (5.76%, 6.59%) | 18575 | 1.07% (1.00%, 1.15%) | 1,730 (1,615; 1,845) |
| Female | 28,459 | 5.75% (5.42%, 6.09%) | 11600 | 0.71% (0.67%, 0.75%) | 1,634 (1,540; 1,728) |
| **Age** |  |  |  |  |  |
| 15-44 | 21,335 | 7.20% (6.73%, 7.66%) | 524 | 0.03% (0.03%, 0.04%) | 1,536 (1,437; 1,635) |
| 45-64 | 14,406 | 6.18% (5.78%, 6.58%) | 4657 | 0.52% (0.49%, 0.55%) | 895 (837; 953) |
| 65-74 | 5,576 | 3.16% (2.67%, 3.66%) | 5663 | 3.13% (2.65%, 3.61%) | 181 (153; 209) |
| 75+ | 4,778 | 3.30% (2.53%, 4.08%) | 19330 | 11.64% (9.22%, 14.06%) | 166 (131; 201) |

<sup>1</sup>All estimates of prevalence adjusted for imperfect test sensitivity and specificity (see text for details). Responses have been re-weighted to account for sample design and for variation in response rate (age, gender, ethnicity and deprivation) in final column to be representative of the England population (18+); <sup>2</sup> Infection fatality ratios were calculated excluding care home residents. Confirmed COVID-19 death counts were obtained from https://fingertips.phe.org.uk/static-reports/mortality-surveillance/excess-mortality-in-england-week-ending-17-Jul-2020.html. Deaths in care homes by age on 12 June 2020 were obtained from https://www.ons.gov.uk/peoplepopulationandcommunity/birthsdeathsandmarriages/deaths/articles/deathsinvolvedcovid19inthecaresectorenglandandwales/deathsoccurringupto12june2020andregisteredupto20june2020provisional. Total deaths in care home residents up to 17 July 2020 were obtained from https://www.ons.gov.uk/peoplepopulationandcommunity/birthsdeathsandmarriages/deaths/datasets/numberofdeathsincarehomesnotifiedtothecarequalitycommissionengland. The age stratified estimates of COVID-19 deaths were estimated using the total deaths from 17 July and the age distribution from 12 June. We assume the age distribution of deaths did not change between 12 June and 17 July 2020.

## Discussion

This is to our knowledge, the largest community-based evaluation of SARS-CoV-2 antibody prevalence, and the only nationwide study based on unsupervised use of LFIA tests at home. It shows an overall prevalence of 6.0% in England, with the epidemic widely dispersed geographically. Overall we estimate that 3.4 million adults had been infected with SARS-CoV-2 virus in England to the end of June 2020, with the majority of people who developed antibodies reporting symptoms during the peak of the epidemic in March and April 2020. As the epidemic took off it became more concentrated in specific groups including Black, Asian and other minority ethnic groups, and essential workers, particularly those working in health and residential social care. While partially attenuated in the adjusted analyses, these factors persisted and reflect a starkly uneven experience of the COVID-19 epidemic across society.

An unequal burden of COVID-19 morbidity and mortality is emerging from many countries including the USA as well as the UK. (26-30) Our study has the advantage of including ethnicity data alongside information about employment, deprivation, household size and other potential confounders. This allows a more nuanced exploration of the underlying mechanisms for these unequal outcomes.(8) In the UK context we suggest that a higher incidence of infection, rather than differences in infection fatality ratios, underpin the observed excess hospitalisations and mortality in minority ethnic groups. Further research is needed to better understand the reasons behind these higher infection rates among non-white populations and the extent to which they reflect underlying structural inequalities, occupational or other factors.(31)

In common with some other studies we found that current smokers have a lower prevalence of SARS-CoV-2 infection than non-smokers.(6, 32) It is unclear whether this relects unmeasured confounding, differential adoption of preventive behaviours, given the known associations of COVID-19 severity with smoking-related co-morbidities, or whether there may be some biological basis. In this regard, the effect of nicotine on angiotensin converting enzyme 2 (ACE2) receptors, a route of viral entry into cells, has been proposed as a potential mechanism.(33)

Our estimated IFR of 0.90% is in line with a recent large study in Spain which reported 0.83% to 1.07%, lower than the IFR described in Italy (2%), and higher than that reported from a German study (0.38%).(34-36) The overall IFR is dependent on the age and sex distribution of infection, and stratified estimates may be more informative. In estimating the IFR, we may have underestimated the number of infected individuals (leading to higher estimates of IFR), as a result of weakened or absent antibody response in some people, and waning antibody over time.(37) We excluded deaths in care home residents since few such residents were included in our community sample. As shown in our sensitivity analysis, this reduced our estimate of the IFR given that, like many countries, England experienced high numbers of cases and deaths in care home residents.(38)

The clinical spectrum of infection is wide, with just under one third (32%) of people with antibodies reporting no symptoms; this proportion was higher in people over 65 years (49%) as also reported for individuals in long-term care.(39) The national prevalence study in Spain reported that 28.5% or 32.7% were asymptomatic depending on the test, (14) similar to our findings overall, although a systematic review of 16 clinical studies puts the figure at 40-50%.(40) The high prevalence of asymptomatic infection is missed in much routine testing which is based mainly on symptomatic individuals.

Our study has a number of limitatons. As in almost all population surveys, our study showed unequal participation, with lower response from ethnic minority groups and people in more deprived areas. We re-weighted the sample to account for differential response, although this may not have overcome unknown participation biases. An important limitation was the exclusion of children for regulatory reasons as the tests were approved for research use in adults only. We used self-administered home LFIA tests as opposed to “gold standard” laboratory tests based on a blood draw. However, we carried out extensive evaluation of the selected LFIA whch showed it to have acceptable performance in terms of both sensitivity and specificity in comparison with the confirmatory laboratory tests.(21) We also took steps to measure and improve usability, including ability to perform and read an LFIA test, through public involvement and evaluation in a national study of 14,000 people.(22)

Use of the LFIA enabled us to obtain antibody tests on large numbers over an 18-day period, without the need for laboratory or health care personnel. Antibodies were strongly associated with clinical history of confirmed or suspected COVID-19, providing face validity. Although there was a theoretical potential for reporting bias as respondents were not blinded to their test results, there was high concordance of self-reported with clinician-read results from the uploaded photographs. Our results closely tracked other indicators of the epidemic curve and we believe that use of home-based self-tests is a sustainable model for community-based prevalence studies in other populations. These could provide reliable estimates of the timing and extent of the epidemic, the groups most at risk, whilst avoiding the biases of surveillance that relies solely on self-referral for testing.

In conclusion, our finding of substantial inequalities in prevalence of SARS-CoV-2 infection by ethnicity and social deprivation runs counter to suggestions that their excess risk is due predominantly to comorbidities or other biological factors. Specifically, the higher risk of infection in minority ethnic groups may explain their increased risk of hospitalisation and mortality from COVID-19.

## Data Availability

The original datasets generated or analysed, or both, during this study are not publicly
available because of governance restrictions and the identifiable nature of the data.

## Contributions

HW and PE designed the study and drafted the manuscript

MW, KECA, JE, CA, LO, RR conducted the analyses

HW, CA, GC, DA, CAD, WB, AD, GC, SR, PE, provided study oversight

AD and PE obtained funding

All authors have reviewed and approved the final manuscript

## Acknowledgements

HW is a NIHR Senior Investigator and acknowledges support from NIHR Biomedical Research Centre of Imperial College NHS Trust, NIHR School of Public Health Research, NIHR Applied Research Collaborative North West London, Wellcome Trust (UNS32973).

GC is supported by an NIHR Professorship. WSB is the Action Medical Research Professor, AD is an NIHR senior investigator and DA is an Emeritus NIHR Senior Investigator.

SR acknowledges support from MRC Centre for Global Infectious Disease Analysis, National Institute for Health Research (NIHR) Health Protection Research Unit (HPRU), Wellcome Trust (200861/Z/16/Z, 200187/Z/15/Z), and Centres for Disease Control and Prevention (US, U01CK0005-01-02)

PE is Director of the MRC Centre for Environment and Health (MR/L01341X/1, MR/SO 19669/1). PE acknowledges support from the NIHR Imperial Biomedical Research Centre and the NIHR HPRUs in Environmental Exposures and Health and Chemical and Radiation Threats and Hazards, the British Heart Foundation Centre for Research Excellence at Imperial College London (RE/18/4/34215) and the UK Dementia Research Institute at Imperial (MC_PC_17114).

We thank key collaborators on this work -- Ipsos MORI: Stephen Finlay, John Kennedy, Kevin Pickering, Duncan Peskett, Sam Clemens and Kelly Beaver; Institute of Global Health Innovation at Imperial College: Gianluca Fontana, Dr Hutan Ashrafian, Sutha Satkunarajah and Lenny Naar; the Patient Experience Research Centre and the REACT Public Advisory Panel; NHS Digital for access to the NHS Register.

